# A machine learning model for prediction of early-onset neonatal sepsis in low- and middle-income countries: Development and validation study

**DOI:** 10.1101/2025.09.20.25335989

**Authors:** Deepika Kainth, Ayushi Gupta, Pradeep Singh, Satya Prakash, Anu Thukral, Ashok Deorari, Mudit Kapoor, Ramesh Agarwal, Tavpritesh Sethi, M Jeeva Sankar

## Abstract

**Objective:** Early-onset sepsis (EOS), which occurs within the first 72 hours of life, can often be fatal for neonates. Machine learning (ML) models demonstrate promise for timely diagnosis. However, current ML models primarily rely on data from high-income countries, which reduces their applicability to low- and middle-income countries (LMICs) that have a higher burden and different disease profiles. We developed an ML model for the timely prediction of culture-proven EOS in LMICs.

**Methods:** We conducted a secondary analysis of the Delhi Neonatal Infection Study (2011-2014) carried out in three level-3 neonatal units in India. We extracted data for inborn neonates suspected of having EOS and excluded cases of culture-negative sepsis. By implementing a dynamic 80:20 (train:test) data split, we employed two feature selection methods—Boruta and Lasso—across 64 variables and applied five machine learning techniques. We aimed to achieve 90% sensitivity to identify the optimal model based on performance metrics. The developed model was integrated into a web application and validated in an external cohort of neonates born between 2015 and 2021.

**Results:** Of 2,924 neonates, 548 (18.7%) had culture-proven sepsis. The mean gestation and birth weight were 35.3 (±3.8) weeks and 2,112 (±754) g, respectively. The Boruta and random forest classifier yielded the best model, which included 28 perinatal-neonatal variables. The sensitivity and specificity of the model were 90.3% and 40.6%, respectively. In external validation (n=147; 26 culture-proven sepsis cases), the model’s sensitivity, specificity, positive predictive value, and negative predictive value were 92.3%, 37.2%, 24.0%, and 95.7%, respectively. The sensitivity was 100% in asymptomatic neonates with only perinatal risk factors for EOS. Using the model could have reduced antibiotic usage from 74.8% to 55.7% (risk difference: -19.1%; 95% CI: -8.3 to -29.7).

**Conclusions:** The ML model demonstrated high sensitivity and acceptable specificity in predicting early-onset sepsis. This prediction model has the potential to assist in the timely and reliable identification of culture-positive sepsis and may serve as a bedside decision support tool in LMICs.

**What is already known on this topic?:**

- Machine-learning models display a good predictive performance for neonatal sepsis prediction.
- Existing models, developed using data from high-income countries, have concerns regarding their generalisability and have not been externally validated.

**What does this study add?:**

- We developed and externally validated a prediction algorithm using a large dataset, prospectively collected variables, and machine learning techniques to predict early-onset neonatal sepsis.
- The model displayed 90.3% sensitivity and 40.6% specificity.

**How this study might affect research, practice or policy?:**

- Our model, incorporated as a computer-based application, can be an excellent clinical aid for enhancing the clinician’s prediction of neonatal sepsis in low- and middle-income countries.

## Introduction

Reliable diagnosis of sepsis and its appropriate management are critical to reducing all-cause and sepsis-related mortality in neonates. [1] Decades of efforts to identify effective predictive and risk-stratification strategies have largely been unsuccessful. Consequently, healthcare providers, cautious of the high case fatality, often resort to the empirical initiation of antibiotics in neonates with suspected sepsis. An additional 15-28 neonates, many of whom may be asymptomatic, receive antibiotics to treat one truly culture-proven neonate. [2] A consequence of this approach is the spiraling multi-drug antimicrobial resistance (AMR) in up to 50-80% of neonates with culture-proven sepsis. [3]

Several risk scores and prediction models have been developed to reliably predict culture-proven sepsis. However, these models have limited diagnostic accuracy and should be regarded as an aid rather than a replacement for diagnostic methods. [4] The only model that has been externally validated as a clinical tool originated in a high-income setting. [5] It is reasonably accurate and has been incorporated into clinical guidelines for managing sepsis. [6,7] However, its applicability to clinical settings in low- and middle-income countries (LMICs) remains uncertain, as colonization with Group B streptococcus (GBS), a necessary feature, is rarely observed in LMICs, where Gram-negative bacteria predominate. Additionally, the baseline EOS risk, which is also required in the model, is nearly 10 times higher in LMICs than in developed countries. Furthermore, the model is only applicable to neonates >35 weeks’ gestation.

Machine learning (ML) has recently been acknowledged as a breakthrough in predictive modeling. Unlike conventional methods, ML identifies complex multidimensional patterns that may surface hours to days before overt manifestation. The key advantage of ML is its ability for continuous performance improvement with repeated data input. A recent systematic review assessing the diagnostic performance of ML models reported impressive results, with a pooled sensitivity of 0.87 (95% CrI 0.75-0.94) and specificity of 0.89 (95% CrI 0.77-0.95) for neonatal sepsis. [8] All included studies, except one, had a high risk of bias, mainly due to the case-control design, small sample sizes, fewer events per predictor, and lack of external validation. Only one study utilized data from LMICs despite 99% of global neonatal deaths occurring in these settings. [9] These concerns are further increased by the fact that many clinical variables are not usually available in LMIC settings. In a typical secondary or tertiary neonatal unit in an LMIC, perinatal variables (such as Apgar score, need for resuscitation, mode of delivery, birth weight, and signs of chorioamnionitis) and details of overt clinical signs (shock, apnea, need for respiratory support, etc.) are generally accessible. However, many variables used as features in existing ML-models—like maternal age, antenatal visits, maternal fever, maternal leukocyte count, duration of prelabor membranes’ rupture, GBS colonization status, need for intrapartum antibiotics, and certain lab parameters (cord pH, blood counts, inflammatory markers)—are not routinely available. Consequently, the validity and generalizability of current ML-models in LMICs remain uncertain. We aimed to create an ML-based model for predicting early-onset sepsis (EOS) using easily obtainable variables in LMICs. We also aimed to develop a user-friendly web application for simple bedside implementation of the model.

## Methods

The model was developed using data from the Delhi Neonatal Infection Study (DeNIS) Collaboration. [10] We carried out the study in six steps: data pre-processing, cohort development, feature selection, model development, internal validation, and prospective validation (Figure 1). Detailed steps are provided in Table 1. The study methods are reported according to the TRIPOD guidelines for reporting prediction modeling studies. [11]

**Figure 1.**
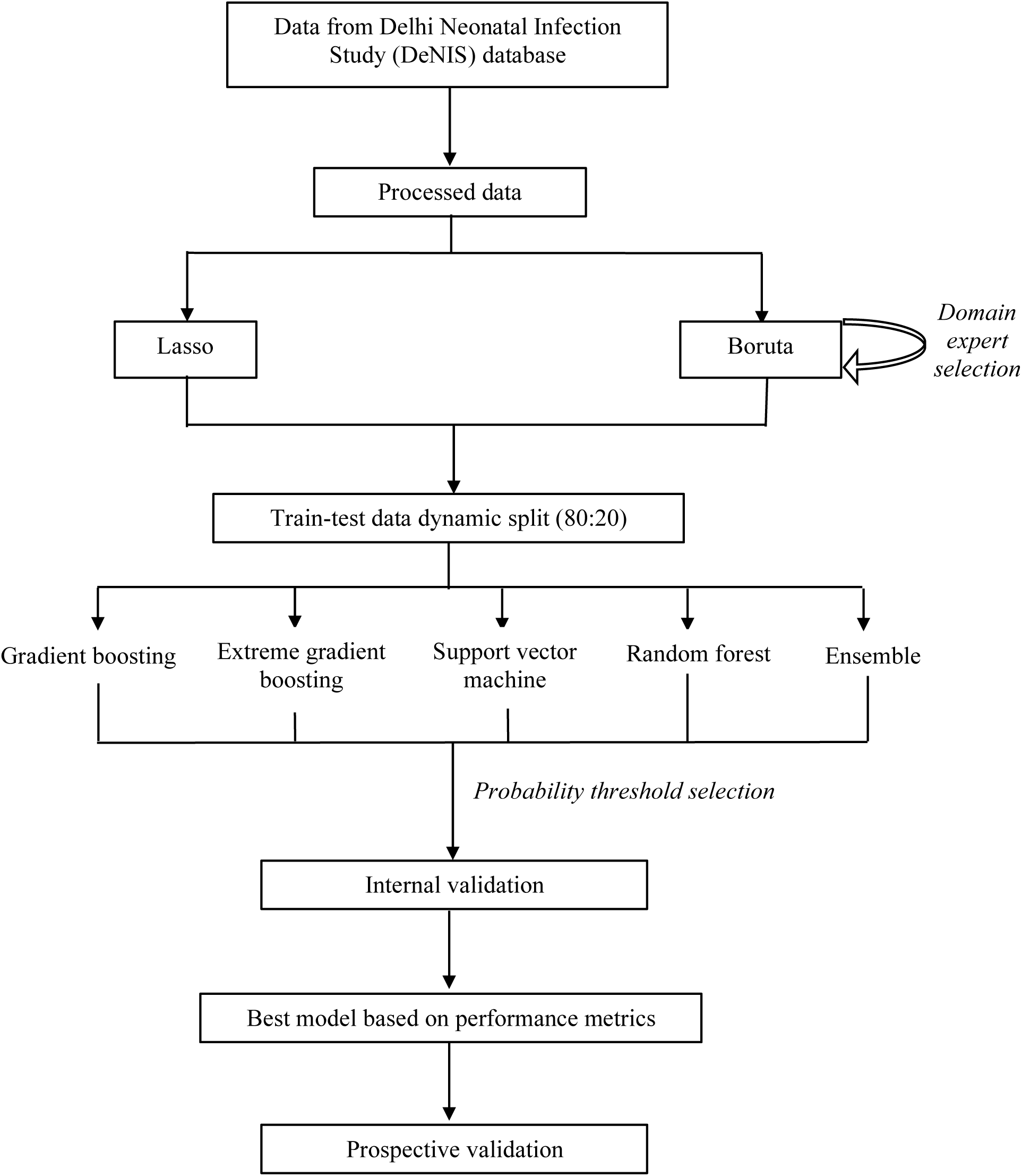
The overall schema of model development and validation.

**Table 1.**
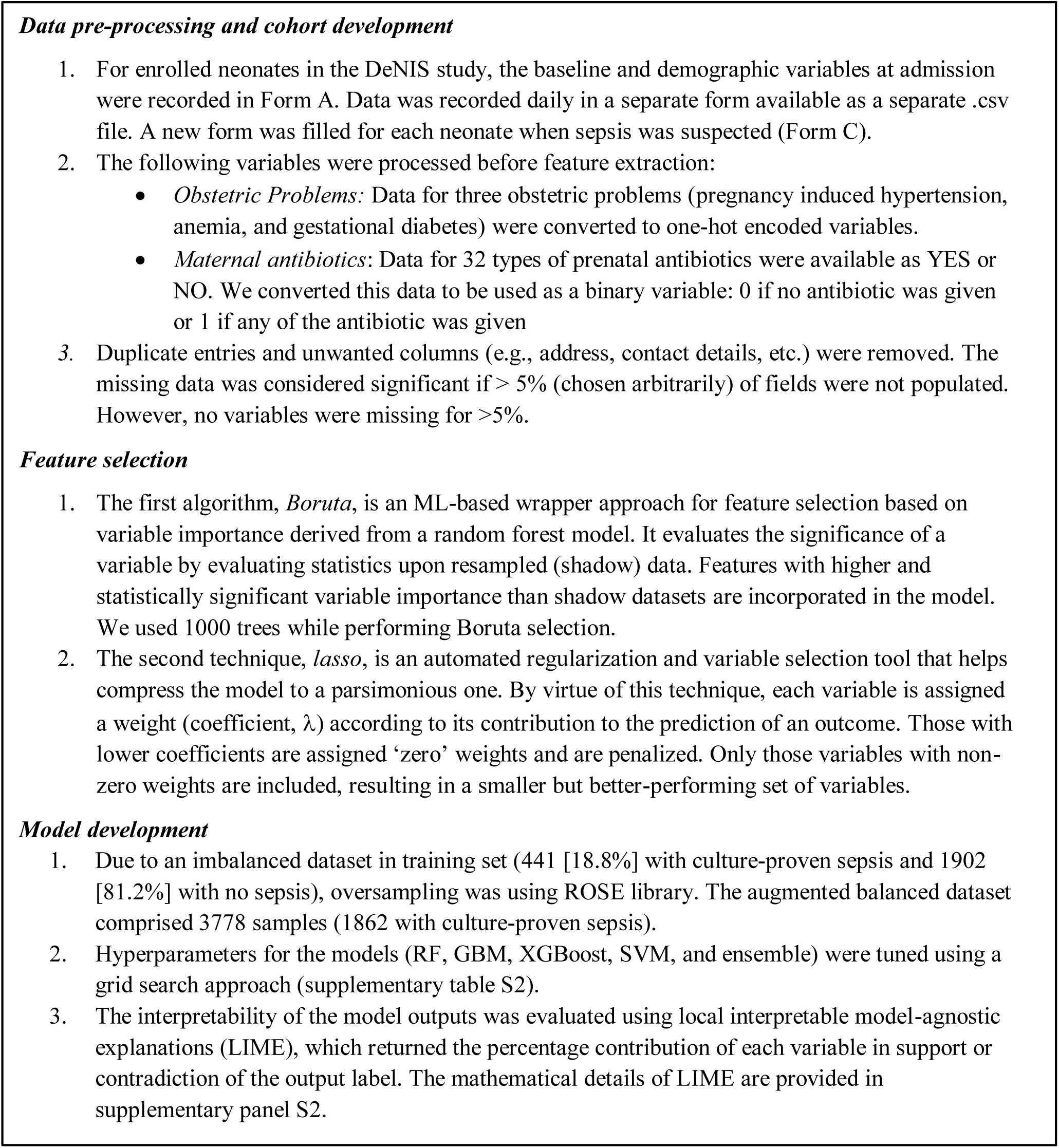
Detailed steps of data pre-processing and model development.

### Source of data and pre-processing

The DeNIS collaboration prospectively enrolled 15417 inborn neonates at three tertiary care centers in New Delhi, India, from March 2011 to October 2014, with published data reporting 13530 neonates enrolled until February 2014. [10] Maternal, perinatal, and neonatal information was collected from all participating centers and recorded on a standardized form during the study. The research staff monitored all enrolled neonates daily for clinical signs of sepsis. Sepsis was suspected in the presence of perinatal risk factors or clinical signs as outlined by the Young Infant Study algorithm. [12] This time point was regarded as the age of suspicion and was recorded in hours of life. The research nurses obtained blood samples for culture from all suspected neonates before initiating antibiotics, which were started at the clinician’s discretion. The final blood culture report was collected 48-72 hours after suspicion. Based on the culture reports and the clinical course, the team of study investigators categorized the neonates as having culture-proven, culture-negative, or no sepsis. Neonates suspected of having sepsis within the first 72 hours of age were eligible for inclusion in this study. For model development, we compared neonates with culture-proven sepsis to those with ‘no sepsis.’ We excluded neonates with culture-negative sepsis (sterile cultures, but the clinical course was consistent with sepsis). The Institutional Ethics Committee at All India Institute of Medical Sciences, New Delhi approved the study protocol. The work was commenced after approval by the committee which serves as the Institutional Review Board. The protocol is registered as IECPG-402/26.05.2022; RT-20/30.06.2022.

### Development and validation cohort

Raw anonymized data from eligible neonates were merged. The train-test data split was performed dynamically, with 80% (training set) exclusively used for feature selection and model development. The remaining 20% (test set) was then utilized for internal validation. For each subsequent data entry in the test data, the probability of sepsis was generated based on the already incorporated data, indicating the model’s dynamic nature and potential for improved performance with increasing data influx. Model performance was assessed using culture-proven sepsis as the reference standard. The experimental steps were conducted in R 3.6.0 using H2O [v.3.36.1.2].

### Feature selection

All maternal and neonatal variables were assessed for inclusion as potential predictors. Maternal characteristics included demographic details, antenatal history, and details of labor. Neonatal variables included birth weight, gestational age, and clinical signs observed at the time of suspicion. The baseline incidence of EOS (expressed as a percentage of NICU admissions) for each site was included to contextualize the model. The need for invasive ventilation, continuous positive airway pressure (CPAP), peripheral intravenous (IV)/arterial cannulation, umbilical catheters, peripherally inserted central catheter (PICC), and medications were classified as ‘care-related variables.’ These variables were considered as inputs in terms of ‘days of exposure’ until at least 24 hours (one visit) before suspicion, ensuring they were potential risk factors rather than initiated due to clinical deterioration. Additionally, the results of the sepsis screen were integrated into the model. The five components of the sepsis screen are outlined in Supplementary Table S1; the screen was considered positive if two of the five components were found to be abnormal.

Supervised feature selection was conducted using two algorithms: Boruta and Least Absolute Shrinkage and Selection Operator (LASSO). [13,14] A pairwise Pearson correlation was performed among the features selected by both methods to ensure the model’s parsimony. Pairs with a coefficient exceeding 0.7 were deemed highly collinear, and only the better-performing variable from each pair was retained.

### Model development

The selected features were used to train classification models utilizing Random Forest (RF), Gradient Boosting Model (GBM), Extreme Gradient Boosting Model (XGBOOST), Support Vector Machine (SVM), and an Ensemble that combines the first four models. Grid search was used to test different combinations of hyperparameters, and the combination yielding the best specificity against a sensitivity of 90% was considered optimal. Results were reported on validation data using these hyperparameters. All five techniques were employed to create models using Boruta and Lasso selection methods (Supplementary table S2).

The primary outcome was the predictive performance of the model, evaluated in terms of area under the receiver operating characteristic (AUROC) curve, sensitivity, specificity, positive predictive value (PPV), and negative predictive value (NPV). Unlike the traditionally chosen probability threshold of 0.5, we examined various probability thresholds ranging from 0.1 to 0.5. Considering the high case fatality rate of neonatal sepsis, our goal was to identify all neonates with sepsis. Thus, we selected a target sensitivity of at least 90% and assessed the corresponding specificity values at each probability threshold for all models. The optimal combination of feature selection technique and model performance was chosen and integrated into an interactive RShiny web application. [15] Using the test set (20% of data), we assessed the discriminatory ability of the best model. We also included a unique interpretability feature in the results for each input data into the model output (Supplementary panel S1). This explainable ML feature helps to explain the prediction of the model in response to a particular input data and improves model transparency, making it more understandable. The complete code in R language, along with the anonymized dataset, is accessible with permission from the authors at https://github.com/tavlab-iiitd/EOS_Prediction_Modeling.

### Prospective validation

All neonates admitted to the neonatal unit at the All India Institute of Medical Sciences, New Delhi, and suspected of sepsis within the first 72 hours of age were included in the prospective validation cohort. Although the unit is one of three study sites for the DeNIS study (development cohort), the data were from a different period and included neonates with a higher level of illness than those in the DeNIS cohort. The model was applied to enrolled neonates, using variables at sepsis suspicion as input to the web-based software developed during the development phase, while maintaining the same evaluation criteria as those used in the development cohort. The cohort was divided into two epochs: *Epoch 1*: Retrospective cohort (January 2015-December 2020) and *Epoch 2*: Prospective cohort (January 2021-September 2021). For Epoch 1, case records of the admitted neonates were retrieved, and data were collected upon suspicion of EOS. For Epoch 2, admitted neonates were prospectively enrolled when sepsis was suspected. Baseline characteristics, clinical signs, and care-related variables at sepsis suspicion were recorded. The final sepsis label was assigned by the clinical team and reviewed independently by two authors, who examined the clinical course and blood culture reports before assigning the label. Performance was evaluated in the overall cohort and in the subgroup of asymptomatic neonates with perinatal risk factors to investigate whether the model demonstrated improved prediction.

### Statistical analysis

We utilized R (version 3.6.0), RStudio (version 1.4.1103), and Stata (version 15.1; StataCorp, College Station, TX) for model development and data analysis. Categorical variables were presented as proportions, while continuous variables were reported as means and standard deviations (SD) or medians (interquartile range), depending on their distribution. The model output was compared with the true disease label, and a 2 × 2 table was constructed to compute sensitivity, specificity, positive and negative predictive values, and likelihood ratios.

### Sample size

We employed the user-defined command *‘pmsampsize’* in Stata to estimate the number of predictor variables that could be identified during the development of new models based on the following assumptions and guidance from Riley et al.: 2,924 neonates, 18.7% outcome prevalence (culture-proven sepsis), a C-statistic of 0.85, events per predictor parameter of 6.0, a shrinkage factor of 0.9, and a small optimism of 0.05. [16] The maximum number of predictors that could be estimated was 91. We employed 67 candidate predictors for model development.

### Patient and public involvement

Patients or public were not involved in the design, or conduct, or reporting, or dissemination of the study.

## Results

The construction of the development cohort is outlined in supplementary figure S1. Of the 3,768 neonates suspected of EOS, 844 were categorized as having culture-negative sepsis and were thus excluded. The data from the remaining 2,924 neonates were analyzed: 548 (18.7%) had culture-proven sepsis, while 2,376 (81.2%) were classified as not having sepsis. The characteristics of the training data, obtained through a dynamic train-test split, are presented in Table 2. The mean birth weight was 2,112 g. Approximately half (51%) were preterm (<37 weeks). Two-thirds presented within 24 hours, with a median (IQR) age at suspicion of 13 (5 to 33) hours. Neonates with culture-proven sepsis had lower gestational age and birth weight. The predominant clinical sign was difficulty breathing (59%), followed by lethargy (46%).

**Table 2:**
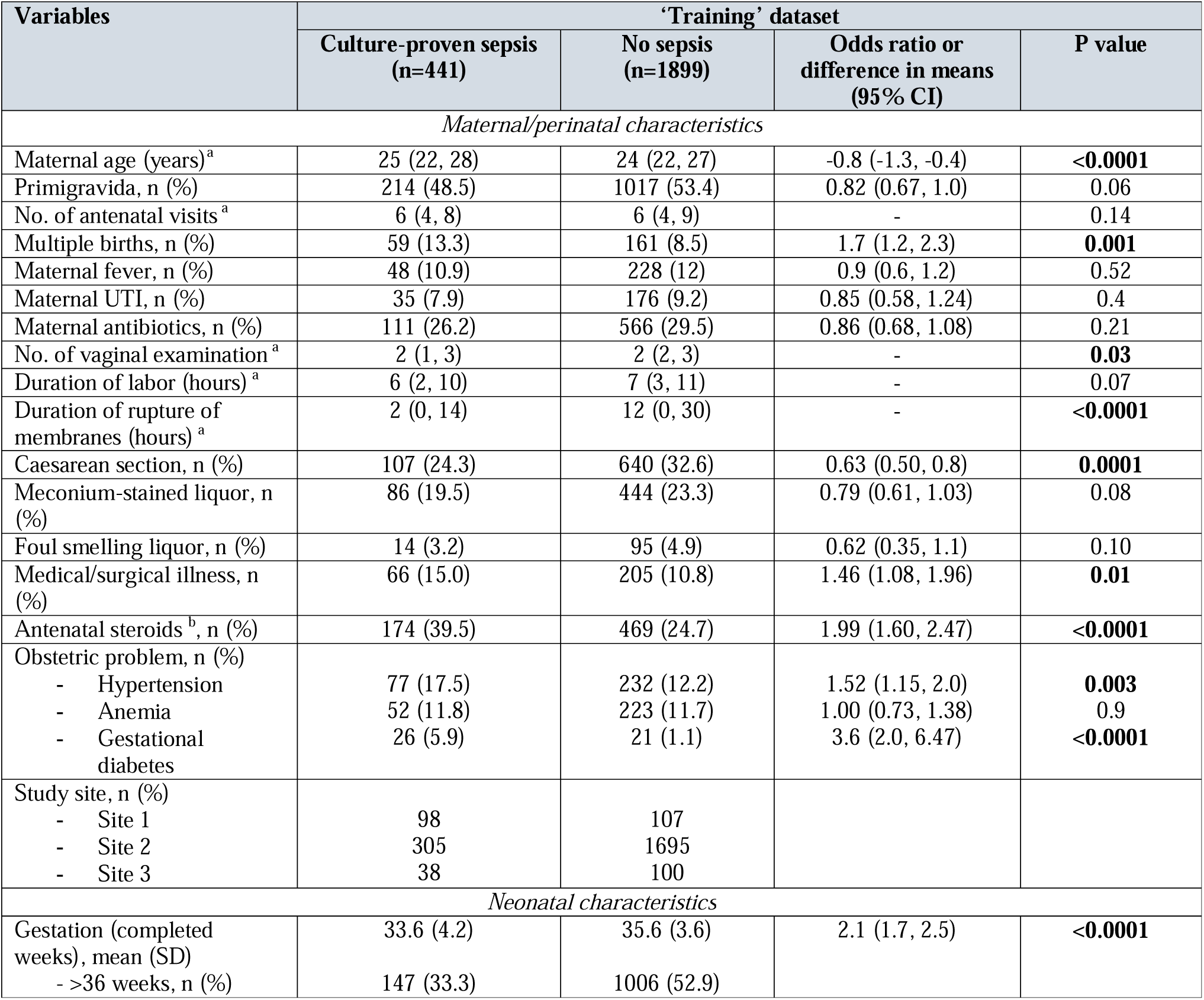
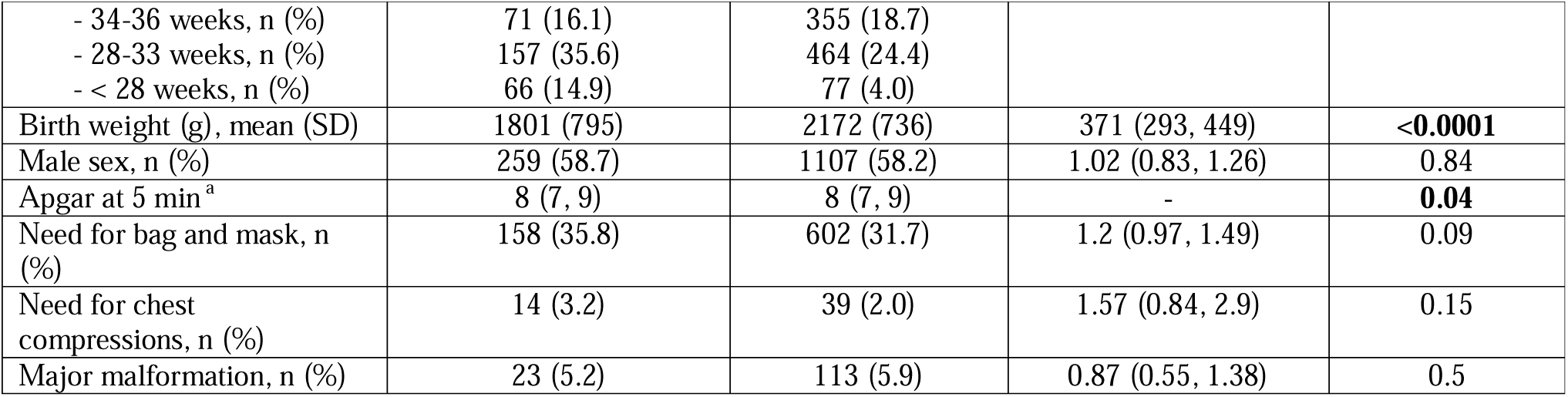
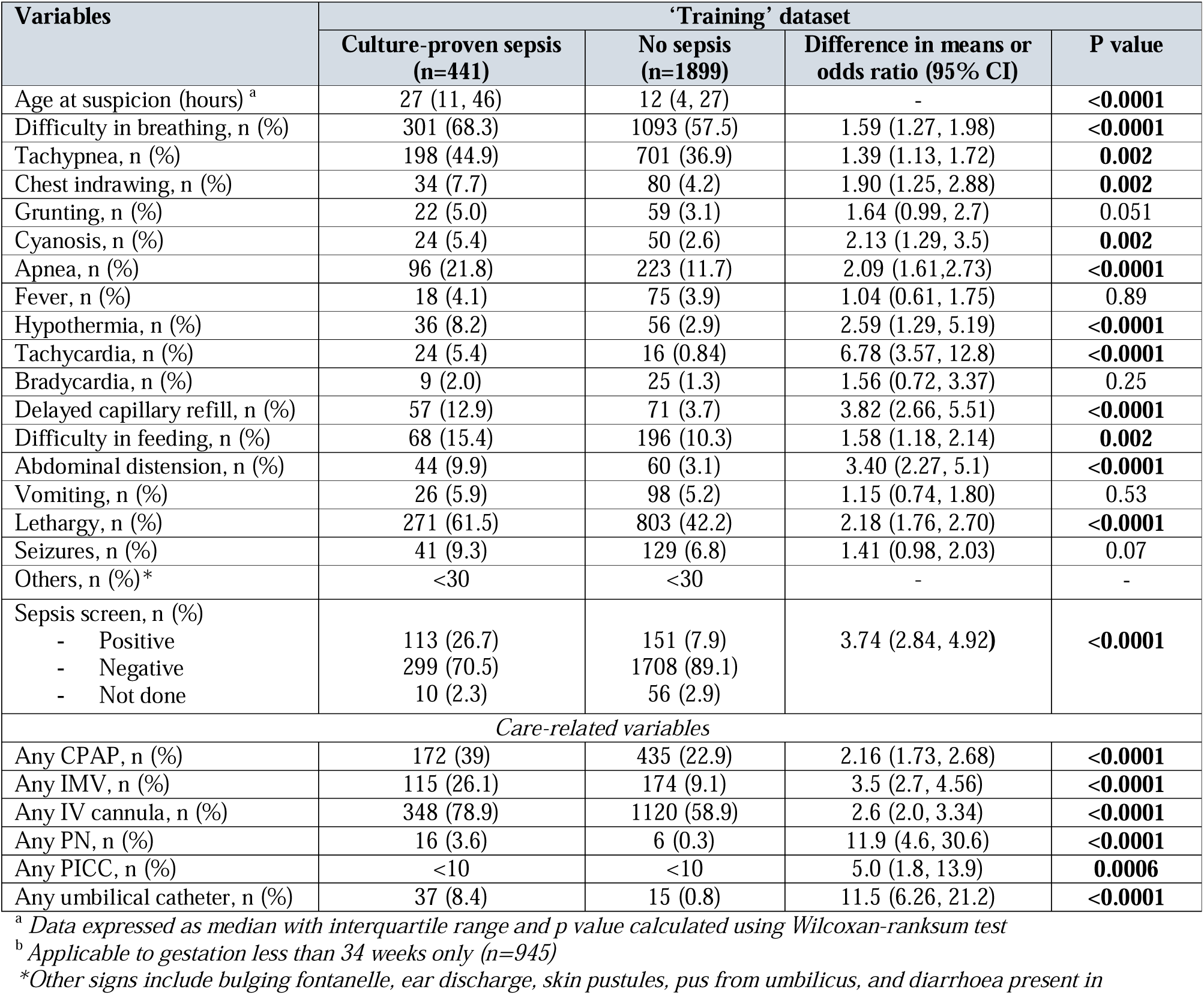
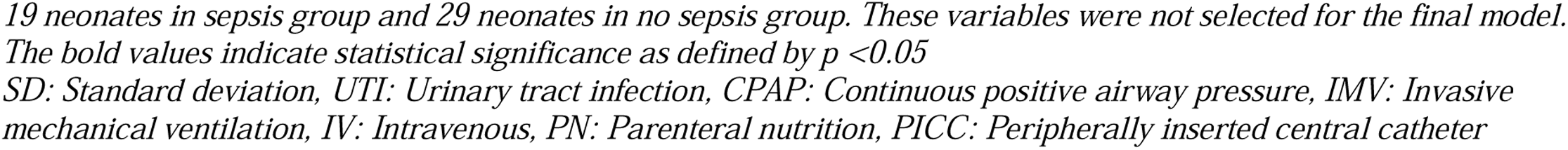
Baseline characteristics of the training cohort used for model development.

| Variables | 'Training' dataset |  |  |  |
| --- | --- | --- | --- | --- |
|  | Culture-proven sepsis<br>(n=441) | No sepsis<br>(n=1899) | Odds ratio or<br>difference in means<br>(95% CI) | P value |
| *Maternal/perinatal characteristics* |  |  |  |  |
| Maternal age (years)<sup>a</sup> | 25 (22, 28) | 24 (22, 27) | -0.8 (-1.3, -0.4) | **<0.0001** |
| Primigravida, n (%) | 214 (48.5) | 1017 (53.4) | 0.82 (0.67, 1.0) | 0.06 |
| No. of antenatal visits<sup>a</sup> | 6 (4, 8) | 6 (4, 9) | - | 0.14 |
| Multiple births, n (%) | 59 (13.3) | 161 (8.5) | 1.7 (1.2, 2.3) | **0.001** |
| Maternal fever, n (%) | 48 (10.9) | 228 (12) | 0.9 (0.6, 1.2) | 0.52 |
| Maternal UTI, n (%) | 35 (7.9) | 176 (9.2) | 0.85 (0.58, 1.24) | 0.4 |
| Maternal antibiotics, n (%) | 111 (26.2) | 566 (29.5) | 0.86 (0.68, 1.08) | 0.21 |
| No. of vaginal examination<sup>a</sup> | 2 (1, 3) | 2 (2, 3) | - | **0.03** |
| Duration of labor (hours)<sup>a</sup> | 6 (2, 10) | 7 (3, 11) | - | 0.07 |
| Duration of rupture of<br>membranes (hours)<sup>a</sup> | 2 (0, 14) | 12 (0, 30) | - | **<0.0001** |
| Caesarean section, n (%) | 107 (24.3) | 640 (32.6) | 0.63 (0.50, 0.8) | **0.0001** |
| Meconium-stained liquor, n<br>(%) | 86 (19.5) | 444 (23.3) | 0.79 (0.61, 1.03) | 0.08 |
| Foul smelling liquor, n (%) | 14 (3.2) | 95 (4.9) | 0.62 (0.35, 1.1) | 0.10 |
| Medical/surgical illness, n<br>(%) | 66 (15.0) | 205 (10.8) | 1.46 (1.08, 1.96) | **0.01** |
| Antenatal steroids<sup>b</sup>, n (%) | 174 (39.5) | 469 (24.7) | 1.99 (1.60, 2.47) | **<0.0001** |
| Obstetric problem, n (%) |  |  |  |  |
| - Hypertension | 77 (17.5) | 232 (12.2) | 1.52 (1.15, 2.0) | **0.003** |
| - Anemia | 52 (11.8) | 223 (11.7) | 1.00 (0.73, 1.38) | 0.9 |
| - Gestational<br>diabetes | 26 (5.9) | 21 (1.1) | 3.6 (2.0, 6.47) | **<0.0001** |
| Study site, n (%) |  |  |  |  |
| - Site 1 | 98 | 107 |  |  |
| - Site 2 | 305 | 1695 |  |  |
| - Site 3 | 38 | 100 |  |  |
| *Neonatal characteristics* |  |  |  |  |
| Gestation (completed<br>weeks), mean (SD) | 33.6 (4.2) | 35.6 (3.6) | 2.1 (1.7, 2.5) | **<0.0001** |
| - >36 weeks, n (%) | 147 (33.3) | 1006 (52.9) |  |  |
| - 34-36 weeks, n (%) | 71 (16.1) | 355 (18.7) |  |  |
| - 28-33 weeks, n (%) | 157 (35.6) | 464 (24.4) |  |  |
| - < 28 weeks, n (%) | 66 (14.9) | 77 (4.0) |  |  |
| Birth weight (g), mean (SD) | 1801 (795) | 2172 (736) | 371 (293, 449) | **<0.0001** |
| Male sex, n (%) | 259 (58.7) | 1107 (58.2) | 1.02 (0.83, 1.26) | 0.84 |
| Apgar at 5 min <sup>a</sup> | 8 (7, 9) | 8 (7, 9) | - | **0.04** |
| Need for bag and mask, n (%) | 158 (35.8) | 602 (31.7) | 1.2 (0.97, 1.49) | 0.09 |
| Need for chest compressions, n (%) | 14 (3.2) | 39 (2.0) | 1.57 (0.84, 2.9) | 0.15 |
| Major malformation, n (%) | 23 (5.2) | 113 (5.9) | 0.87 (0.55, 1.38) | 0.5 |

**Table 2:** Baseline characteristics of the training cohort used for model development (continued)
| Variables | ‘Training’ dataset |  |  |  |
| --- | --- | --- | --- | --- |
|  | Culture-proven sepsis (n=441) | No sepsis (n=1899) | Difference in means or odds ratio (95% CI) | P value |
| Age at suspicion (hours) <sup>a</sup> | 27 (11, 46) | 12 (4, 27) | - | <0.0001 |
| Difficulty in breathing, n (%) | 301 (68.3) | 1093 (57.5) | 1.59 (1.27, 1.98) | <0.0001 |
| Tachypnea, n (%) | 198 (44.9) | 701 (36.9) | 1.39 (1.13, 1.72) | 0.002 |
| Chest indrawing, n (%) | 34 (7.7) | 80 (4.2) | 1.90 (1.25, 2.88) | 0.002 |
| Grunting, n (%) | 22 (5.0) | 59 (3.1) | 1.64 (0.99, 2.7) | 0.051 |
| Cyanosis, n (%) | 24 (5.4) | 50 (2.6) | 2.13 (1.29, 3.5) | 0.002 |
| Apnea, n (%) | 96 (21.8) | 223 (11.7) | 2.09 (1.61,2.73) | <0.0001 |
| Fever, n (%) | 18 (4.1) | 75 (3.9) | 1.04 (0.61, 1.75) | 0.89 |
| Hypothermia, n (%) | 36 (8.2) | 56 (2.9) | 2.59 (1.29, 5.19) | <0.0001 |
| Tachycardia, n (%) | 24 (5.4) | 16 (0.84) | 6.78 (3.57, 12.8) | <0.0001 |
| Bradycardia, n (%) | 9 (2.0) | 25 (1.3) | 1.56 (0.72, 3.37) | 0.25 |
| Delayed capillary refill, n (%) | 57 (12.9) | 71 (3.7) | 3.82 (2.66, 5.51) | <0.0001 |
| Difficulty in feeding, n (%) | 68 (15.4) | 196 (10.3) | 1.58 (1.18, 2.14) | 0.002 |
| Abdominal distension, n (%) | 44 (9.9) | 60 (3.1) | 3.40 (2.27, 5.1) | <0.0001 |
| Vomiting, n (%) | 26 (5.9) | 98 (5.2) | 1.15 (0.74, 1.80) | 0.53 |
| Lethargy, n (%) | 271 (61.5) | 803 (42.2) | 2.18 (1.76, 2.70) | <0.0001 |
| Seizures, n (%) | 41 (9.3) | 129 (6.8) | 1.41 (0.98, 2.03) | 0.07 |
| Others, n (%)* | <30 | <30 | - | - |
| Sepsis screen, n (%) |  |  |  |  |
| - Positive | 113 (26.7) | 151 (7.9) | 3.74 (2.84, 4.92) | <0.0001 |
| - Negative | 299 (70.5) | 1708 (89.1) |  |  |
| - Not done | 10 (2.3) | 56 (2.9) |  |  |
| Care-related variables |  |  |  |  |
| Any CPAP, n (%) | 172 (39) | 435 (22.9) | 2.16 (1.73, 2.68) | <0.0001 |
| Any IMV, n (%) | 115 (26.1) | 174 (9.1) | 3.5 (2.7, 4.56) | <0.0001 |
| Any IV cannula, n (%) | 348 (78.9) | 1120 (58.9) | 2.6 (2.0, 3.34) | <0.0001 |
| Any PN, n (%) | 16 (3.6) | 6 (0.3) | 11.9 (4.6, 30.6) | <0.0001 |
| Any PICC, n (%) | <10 | <10 | 5.0 (1.8, 13.9) | 0.0006 |
| Any umbilical catheter, n (%) | 37 (8.4) | 15 (0.8) | 11.5 (6.26, 21.2) | <0.0001 |

<sup>a</sup> Data expressed as median with interquartile range and p value calculated using Wilcoxon-ranksum test
<sup>b</sup> Applicable to gestation less than 34 weeks only (n=945)
<sup>\*</sup>Other signs include bulging fontanelle, ear discharge, skin pustules, pus from umbilicus, and diarrhoea present in
19 neonates in sepsis group and 29 neonates in no sepsis group. These variables were not selected for the final model. The bold values indicate statistical significance as defined by $p < 0.05$ SD: Standard deviation, UTI: Urinary tract infection, CPAP: Continuous positive airway pressure, IMV: Invasive mechanical ventilation, IV: Intravenous, PN: Parenteral nutrition, PICC: Peripherally inserted central catheter

Feature selection was conducted on 67 variables categorized as maternal, neonatal, and care-related (supplementary table S3). We also included ‘centre’ as an input due to the considerable variation in the prevalence of culture-proven EOS at each site (site 1: 8.8%; site 2: 4.9%; and site 3: 2.1%); this was expressed as the percentage of admitted neonates with EOS. The correlation matrix among the candidate predictors is shown in supplementary figure S2. Variables that were strongly indicative of sepsis (e.g., pus from the umbilicus) and those lacking biological plausibility for predicting sepsis were excluded. The Boruta technique proved to be the most effective, yielding a high-performing set of 38 variables (figure 2). Further filtering based on Pearson correlation among Boruta-selected features resulted in 28 variables, including 6 maternal, 4 neonatal, 12 clinical signs, and 6 care-related variables (Table 3). The top three variables identified were age at suspicion, sepsis screen, and birth weight. The feature set selected using Lasso is detailed in supplementary table S4.

**Figure 2.**
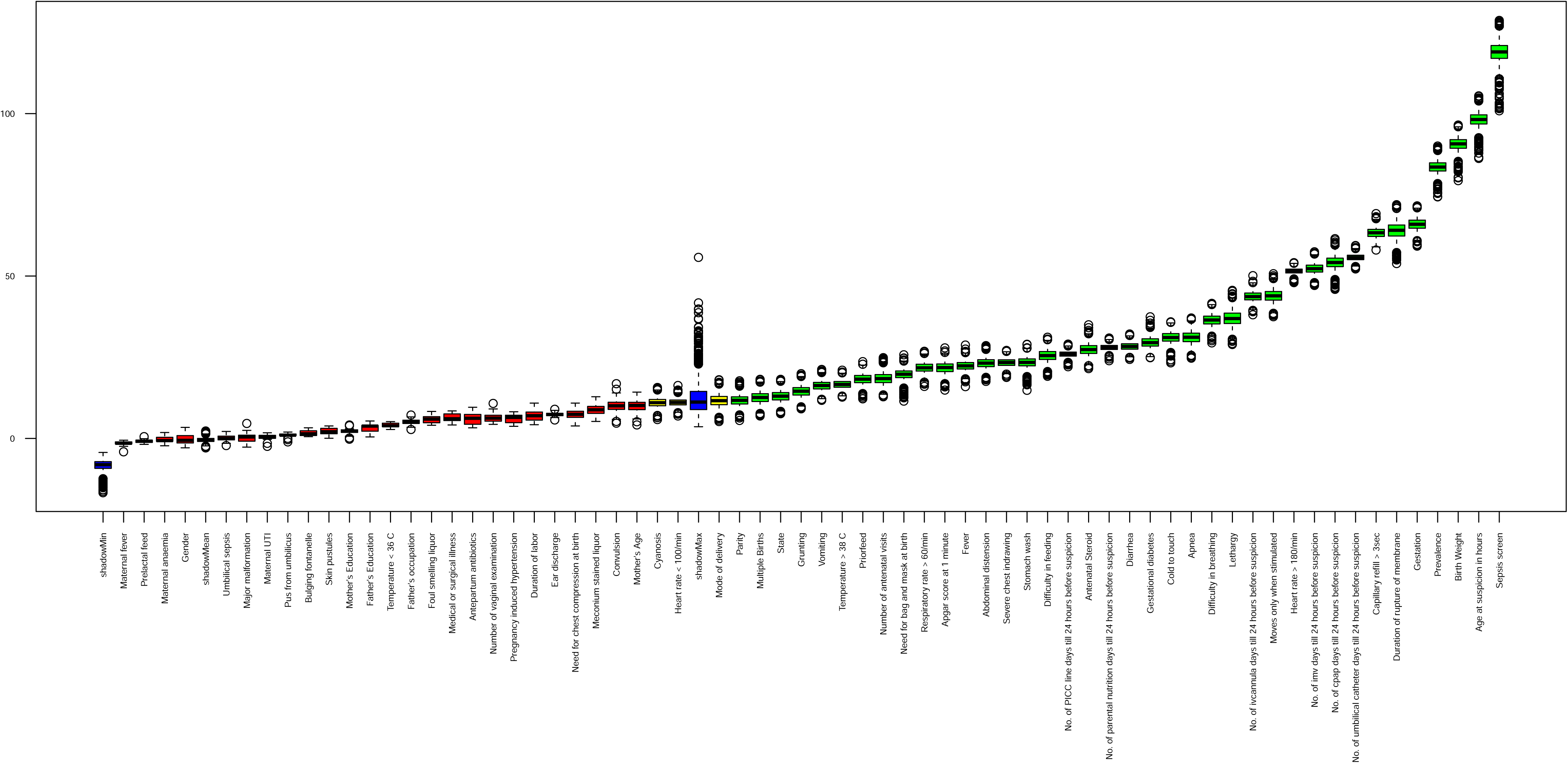
Feature selection using Boruta. *Caption:* The performance of the ‘shadow’ variables is depicted as their maximum Z score (shadowMax), minimum Z score (shadowMin), and mean Z score (shadowMean) (blue). Those variables with Z score higher than shadowMax are depicted in order of their increasing predictive importance (Red). These variables were selected in the model. Variables with a Z score lower than shadowMax (Yellow) decreased the performance of the model and were not selected by Boruta during feature selection

**Table 3:** Final set of maternal, neonatal, and care-related variables.

| Maternal characteristics | Neonatal characteristics | Clinical features |
| --- | --- | --- |
| • Multiple gestation<br>• Gestational diabetes<br>• Number of antenatal visits<br>• Parity<br>• Antenatal steroids<br>• Duration of rupture of membranes (hours)* | • Gestational age*<br>• Birth weight*<br>• Apgar at 1 minute<br>• Age at suspicion of sepsis (hours)*<br>**Care-related variables:\***<br>• Days of CPAP<br>• Days of mechanical ventilation<br>• Days of IV cannula<br>• Days of PICC line<br>• Days of umbilical catheter<br>• Days of parenteral nutrition | • Feeding difficulty<br>• Vomiting<br>• Abdominal distension<br>• Breathing difficulty<br>• Apnoea<br>• Respiratory rate $> 60/\text{min}$<br>• Fever<br>• Cold to touch<br>• Heart rate $> 180/\text{min}$ or $< 100/\text{min}$<br>• Capillary refill $> 3 \text{ sec}$<br>• Lethargy<br>• Sepsis screen |

\*Expressed as continuous data
CPAP: Continuous positive airway pressure; IV: Intravenous; PICC: Peripherally inserted central catheter

### Model performance on the internal validation dataset

Of the 584 neonates in the test set, 107 had culture-proven sepsis (see supplementary table S5). With a target sensitivity of 90% for the best-performing model, RF achieved a sensitivity of 90.2%, specificity of 40.6%, PPV of 25.3%, and an NPV of 95.1% at a probability threshold of 0.33 (refer to supplementary figure S3, table S6, and table S7). The performance of models developed using Lasso is detailed in supplementary table S8. In the test set, 302 neonates (51.7%) were started on antibiotics at the discretion of clinicians. Utilizing our model could have reduced antibiotic usage from 51.7% to 46.9% (n=259). In contrast, 8 of the 91 neonates with culture-proven sepsis would have been misclassified as not having sepsis.

The model was incorporated into a web-based R Shiny application, ‘Neonatal Sepsis Suite,’ which was subsequently used for prospective validation (http://103.25.231.39:3838/NeonatalSepsisSuiteModular/<u>#</u>). After entering the input variables upon suspicion, the output is displayed as ‘Positive’ or ‘Negative.’ If any input variable(s) are unavailable, the probability is generated based on the other available variables.

### Prospective validation

A total of 308 neonates were prospectively enrolled, of whom 161 with culture-negative sepsis were excluded. Of the remaining 147 neonates (126 neonates in retrospective and 21 neonates in prospective cohort), 26 had culture-proven sepsis, while 121 (39.3%) were classified as having no sepsis. The characteristics of the prospective validation cohort are provided in Supplementary Table S9. Compared to the development cohort, neonates in the validation phase were more premature (31 weeks vs. 35 weeks), had a lower birth weight (1574 g vs. 2112 g), and were predominantly born via cesarean section (54% vs. 32%). The proportion requiring invasive lines (16% vs. 2%) and parenteral nutrition (9.5% vs. 0.6%) was also higher in the validation cohort. The most common clinical signs were respiratory (tachypnea, increased oxygen requirement) and gastrointestinal (abdominal distension, vomiting). Overall, the model achieved 92.3% sensitivity, 37.2% specificity, 24.0% PPV, and 95.7% NPV in the validation cohort (supplementary table S10). A total of 110 (74.8%) neonates were started on antibiotics based on the clinician’s discretion. Our model could have potentially reduced antibiotic usage from 74.8% to 55.7% (82/147; risk difference: -19.1%; 95% CI: -8.3 to -29.7), as 28 additional neonates would have been accurately classified as ‘no sepsis.’ Further, two neonates with culture-proven sepsis would have been misclassified as having no sepsis. One of them was born at 29 weeks with a birth weight of 1560 g to a mother with prolonged rupture of membranes (25 h) and developed apnea and respiratory distress at 6 hours of age. The second neonate (40 weeks; 2502 g) presented at 4 hours of life with prolonged capillary refill time, tachycardia, and respiratory distress. Despite the model output indicating ‘no sepsis,’ the clinician’s discretion would likely have overruled the decision regarding antibiotics in both cases.

The sensitivity and specificity were 95.4% and 36.5%, respectively, in preterm neonates (n=126; 22 culture-proven sepsis) and 75% and 37.5%, respectively, in term neonates (n=21; 4 culture-proven sepsis). Further, in those with normal birth weight (n=69; 12 culture-proven sepsis), the sensitivity was 83.3% and specificity was 66.7%, compared to 100% sensitivity and 26.6% specificity in low birth weight neonates (n=78; 14 culture-proven sepsis).

47 (31.5%) neonates were asymptomatic and evaluated for EOS only based on perinatal risk factors at birth. Among them, 3 had culture-proven sepsis, which was accurately identified by the model; 23 without sepsis were also correctly classified by the model, leading to 100% sensitivity (Supplementary table S11). Of the 47 neonates, 36 were started on antibiotics based solely on risk factors. Using our model would have accurately classified 15 (41.7%) of these neonates as having no sepsis, thereby avoiding the empirical initiation of antibiotics. The model would not have missed any cases of culture-proven sepsis in these neonates. Supplementary panel S2 summarizes the case scenarios of three neonates enrolled in the validation phase, including their clinical features at the time of sepsis suspicion, the model output, and the interpretability for the label assigned by the model based on the features.

The supplementary content is available separately as ‘Supplementary material.

## Discussion

Our ML-based model for predicting early-onset sepsis (EOS) demonstrated a high sensitivity of 90.2% and specificity of 40.6% in the development cohort, as well as a sensitivity of 90.3% and NPV of 94.2%, along with reasonable specificity of 40.5% in the external validation cohort. Implementing our model could have resulted in approximately a 20% reduction in antibiotic usage. The model’s high sensitivity can help clinicians at the bedside rule out culture-positive sepsis with reasonable confidence, particularly in asymptomatic neonates with perinatal risk factors for sepsis, as all neonates were accurately classified as not having sepsis.

The prediction models available in literature were examined in a recent systematic review, where 36 clinical decision support algorithms were developed that utilized non-invasive clinical parameters. Of these, ten deployed ML techniques and focused on continuously monitored vital signs. There was considerable variability due to methodological issues, hindering the pooling of results. Additionally, another systematic review analysed 19 studies that developed ML models using routinely collected clinical data. [17] Of these, four specifically created ML models for EOS using data only from high-income countries. Although these models achieved a pooled AUROC of 0.98, they were deemed to carry a high risk of bias due to small sample sizes, low events per predictor, and a lack of external validation. In contrast, our model was developed using a large sample size using a multicentre cohort, a high event rate, a large number of clinical predictors with automated feature selection, and external validation, placing our study at a low risk of bias and applicability. We utilized automated feature selection techniques involving maternal, neonatal, and care-related variables. Boruta, which includes an in-built RF classifier, can handle both tree-based and non-tree-based algorithms and multi-dimensional interactions. Although computationally intensive, it provides robustness and stability in noisy datasets. [18] In our study, Boruta exhibited the best diagnostic performance, resulting in the selection of 28 variables. Conventionally recognized predictors, such as birth weight, gestation, and duration of rupture of membranes, were among the most significant. [19,20] The presence of maternal fever and receipt of intrapartum antibiotics were not selected, unlike the widely used EOS calculator, which includes them in its five most important predictors. [21] This indicates the importance of horizontal transmission of infection in early-onset sepsis within LMIC settings. Among the clinical signs, tachycardia, lethargy, and prolonged capillary refill time were significant, similar to findings in other models.

Despite the best efforts to improve model performance, specificity remained low. This is likely due to targeting high sensitivity given the high fatality of EOS, where we intended to capture (and not miss) all truly diseased neonates. This enhanced the model’s performance as a screening tool at the expense of low specificity. Furthermore, low specificity in the context of low disease prevalence (18.1%) likely led to a low PPV (25.3%) for our model. Three out of four neonates without culture-proven sepsis were incorrectly labeled as having sepsis. This phenomenon was also observed in models developed by Masino et al., which reported a low PPV (23%) for sepsis prediction. [23] To address this concern, Repetto et al. utilized balancing techniques, similar to our study, which minimized skewness and improved PPV (82%).[22] However, their sample size was small (n=236). In contrast, the model developed by Stocker et al. using RF targeted higher specificity (90%) and consequently had low sensitivity (61%). The PPV was very low (9.8%), possibly due to low prevalence (1.7%). [24] The PPV of our model may improve in settings with a high prevalence of culture-proven sepsis.

Our study’s strengths include a large sample size that reduces model variance and the risk of overfitting. The use of automated feature selection is superior to the conventional approach, where manual selection and elimination of known variables are based on literature searches and traditional statistical methods. The utility of the latter technique is limited by its inability to identify multicollinearity and complex interactions among variables. Our model is a straightforward bedside-inclusive model applicable to neonates of all gestations and birth weights. Incorporating our model into a bedside application that provides a predicted probability of sepsis within minutes could facilitate its widespread and efficient implementation. The automated feature selection methodology could offer a platform and serve as a guide for developing more context-specific ML models in neonatal units at various levels of healthcare. Furthermore, a unique feature of our ML model is its dynamic nature and ability to learn and improve based on new incoming data, enhancing its performance with increased use.

Our study has several limitations. First, using our model may lead to overtreatment due to its low PPV. Although we intended to prioritize high sensitivity, a low area under the curve (AUC) of 0.65 and specificity of 54% might not provide a bedside physician with sufficient confidence to utilize the model. Our model also missed two (7.7%) neonates with culture-proven sepsis. Second, we excluded neonates with culture-negative sepsis, who constituted nearly 25% of those suspected of having sepsis. This group represents a poorly defined population, and their inclusion could have raised concerns regarding the misclassification of neonates without sepsis as having sepsis. Additionally, our model was primarily developed to predict the occurrence of culture-proven sepsis to limit antibiotic use. The Early Onset Sepsis (EOS) calculator, which is widely utilized in developed countries, was also created to identify culture-proven cases. [21] Whether culture-negative sepsis cases were included in the model’s development remains unclear. Therefore, it was essential to include only those with culture-positive sepsis and compare them to those classified as ‘no sepsis.’ Third, our model necessitates 29 input variables, including those with very low prevalence (e.g., PICC lines, umbilical catheters) and laboratory parameters (sepsis screen/CRP), which makes our model less parsimonious and requires additional laboratory investigations. Furthermore, variables such as PICC line days and number of antenatal visits are often unavailable in most secondary and tertiary centers, especially those serving outborn neonates. Lastly, the validation cohort was probably not an ideal choice for external validation, as the neonates were from one of the study sites included in the development cohort. Additionally, a large proportion of neonates belonged to the retrospective period of the ambispective cohort used for validation. However, the cohort was temporally distinct and included sicker neonates compared to the development cohort. Despite these limitations, our model is the first of its kind from LMICs and can guide early prediction of sepsis. To address the limitations, we plan to validate our model externally at diverse secondary and tertiary centers. Additionally, to evaluate the true clinical usefulness of our model for bedside use, a randomized controlled trial comparing the effects of decision-making based on the ML model with that of clinical decision-making in Indian settings. We also intend to expand its application to include neonates with culture-negative sepsis, aiming to generalize our model for all neonates with suspected sepsis.

## Conclusions

Our ML model demonstrated 90.3% sensitivity and reasonably good specificity of 40.6% for predicting culture-proven EOS at a level-3 neonatal care unit. This tool has the potential to serve as a useful bedside decision support tool in timely and reliable prediction of culture-proven sepsis in low- and middle-income countries.

## Data Availability

Deidentified participant data used for secondary analysis is available with permission from the authors at https://github.com/tavlab-iiitd/EOS_Prediction_Modeling.

## Authors’ contribution

DK, RA, and MJS conceptualized the study. AT, RA, and AD provided critical input in the conceptualization and design of the study. DK, AG, and SP collected the data and drafted the initial manuscript. DK, AG, and PS analyzed the data and developed the model. TS and MK supervised the analysis and technical aspects of the study. MJS supervised the collection, statistical analysis, and interpretation of data. DK, AG, PS, SP, AT, RA, MK, AD, TS, and MJS critically reviewed and revised the manuscript for important intellectual content and approved the final manuscript as submitted. Dr. M Jeeva Sankar will serve as the guarantor for this submission.

## Acknowledgments

We acknowledge the investigators of the Delhi Neonatal Infection Study (DeNIS) collaboration: Kailash Chandra Aggarwal, M S Prasad, Harish Chellani, Manorama Deb, Rajni Gaind, Sugandha Arya, Sumita Saluja, Siddarth Ramji, S K Prakash, Surinder Kumar, Manoj Modi, Neeraj Gupta, Ashish Jain, Vinod K Paul, Arti Kapil, Suman Chaurasia, Purva Mathur, and Manju Saksena. We also acknowledge the Indian Council of Medical Research (ICMR) for generously funding the Centre for Advanced Research at AIIMS, New Delhi, that coordinated the DeNIS collaboration.

TS also acknowledges support from the Wellcome Trust/DBT India Alliance Fellowship IA/CPHE/501504.

