## Supplemental Data 1 for "A machine learning model for prediction of early-onset neonatal sepsis in low- and middle-income countries: Development and validation study"

**Supplementary Material**

| **S. No.** | **Content** | **Page No.** |
| --- | --- | --- |
| **1.** | **Supplementary table S1.** Components of sepsis screen | 2 |
| **2.** | **Supplementary table S2.** Model parameters used for machine learning techniques | 3 |
| **3.** | **Supplementary panel S1.** LIME algorithm for interpretability of data | 4 |
| **4.** | **Supplementary figure S1.** Construction of the cohort for model development | 5 |
| **5.** | **Supplementary table S3**. Maternal, neonatal and care-related variables screened for feature selection (67) | 6 |
| **6.** | **Supplementary figure S2**. Correlation matrix of variables | 7 |
| **7.** | **Supplementary table S4.** List of features selected using lasso feature selection | 8 |
| **8.** | **Supplementary table S5.** Comparison of characteristics of the test data and training data during model development phase | 9 |
| **9.** | **Supplementary figure S3.** Performance of the RF model at various probability thresholds from 0 to 0.5 | 10 |
| **10.** | **Supplementary table S6.** Performance metrics of machine learning model using Boruta feature selection | 11 |
| **11.** | **Supplementary table S7.** 2x2 table depicting the performance of the Random Forest model in the development phase | 12 |
| **12.** | **Supplementary table S8.** Performance of models developed using lasso feature selection | 13 |
| **13.** | **Supplementary table S9:** Baseline characteristics of neonates enrolled in the validation phase | 14 |
| **14.** | **Supplementary table S10.** 2x2 table depicting the performance of the Random Forest model in the validation phase | 15 |
| **15.** | **Supplementary table S11.** 2x2 table depicting the performance of the model in the validation phase for asymptomatic neonates with perinatal risk factors | 16 |
| **16.** | **Supplementary panel S2.** Case-based scenarios from the validation phase | 16-17 |

**Supplementary table S1. Components of sepsis screen***

| **Component** | **Abnormal value** |
| --- | --- |
| Absolute neutrophil count | Low counts as per Manroe chart for term and Mouzinho’s chart for VLBW infants |
| Total leukocyte count | < 5000/mm^3^ |
| Immature/total neutrophil ratio | > 0.2 |
| Micro-Erythrocyte sedimentation rate | >15 mm fall in 1^st^ hour |
| C reactive protein (CRP) | >1 mg/L |

********All the components were not uniformly available across the study sites in the development phase. Components like immature/total neutrophil ratio are not based on automated cell counters and are therefore operator-dependent. Further, data could not be extracted for individual components and*

*the results were available as ‘positive’ or ‘negative’.*

**Supplementary table S2. Model parameters used for machine learning techniques using grid search approach**

| **Hyperparameters** | **Range used for training the model** |
| --- | --- |
| Trees | 10 to 250 at an interval of 10 |
| Maximum depth | 1 to 20 at an interval of 1 |
| Threshold | 0.1 to 0.5 at an interval of 0.01 |
| ***Model parameters selected for the final model**** | |
| **Machine learning model** | **Parameters used in model training** |
| Random Forest | nfolds = 10, ntrees = 120, mtry = 5, max_depth = 8 |
| Gradient Boosting Model | nfolds = 10, ntrees = 10, learn_rate = 0.1 |
| Extreme Gradient Boosting Model | nfolds = 10, ntrees = 10, learn_rate = 0.1 |
| Support Vector Machine Learning | gamma = -1, rank_ratio = -1 |

**All possible combinations of hyperparameters were analysed and the combination that resulted in highest specificity at 90%*

*sensitivity was used as the final model (depicted here for each model)*

**Supplementary panel S1. LIME algorithm for interpretability of data**

Mathematically, local surrogate models (LIME) with interpretability constraint can be expressed as follows:

explanation(x) = argmin_g∈G_ L(f,g,π_x_) + Ω(g)

The explanation model for instance ‘x’ is the model ‘g’ (e.g., linear regression model) that minimizes loss ‘L’ (e.g., mean squared error), which measures how close the explanation is to the prediction of the original model ‘f’ (e.g., an xgboost model). In contrast, the model complexity ‘Ω(g)’ is kept low (to prefer fewer features). ‘G’ is the family of possible explanations, for instance, all possible linear regression models. The proximity measure ‘πx ’ defines how large the neighborhood around instance ‘x’ is that we consider for the explanation. In practice, LIME only optimizes the loss part. The user must determine the complexity, e.g., by selecting the maximum number of features the linear regression model would use.

**Supplementary figure S1. Construction of the cohort for model development**

**4664** neonates with first episode of sepsis suspicion

**2924** neonates with early-onset sepsis

**548** culture positive sepsis

**2376**

no sepsis

**15417** inborn neonates admitted to 3 neonatal units in New Delhi

(2011-2014)

**4965** neonates evaluated for sepsis

(5308 episodes)

**3768** neonates included for the first episode

**896** neonates with first episode beyond 72 h

of life excluded

**10452** neonates never evaluated for sepsis

throughout the hospital admission

**301** neonates excluded for multiple

episodes of sepsis suspicion

**844** labelled as culture-negative

clinical sepsis excluded

**Supplementary table S3. Maternal, neonatal and care-related variables screened for feature selection (n=67)**

| **Maternal characteristics** | **Neonatal characteristics** | **Clinical features** |
| --- | --- | --- |
| Study site  Maternal age  Mother’s education  Father’s education  Father’s occupation  State of residence  Place of delivery*  Parity  Multiple gestation  Diabetes/Hypertension/  Anemia  Medical or surgical illness  Number of antenatal visits  Antenatal steroids  Maternal fever  Urinary tract infection  Use of antibiotics before  delivery  Duration of labor  Duration of rupture of  membranes  Number of vaginal  examinations  Mode of delivery  Foul smelling liquor | Gestational age  Birth weight  Weight in NICU*  Gender  Cry at birth*  Need for bag and mask  Need for chest compressions  Apgar at 1 min  Apgar at 5 min  Meconium-stained liquor  Major malformation  Age at suspicion of sepsis    ***Care related variables:***  Pre-lacteal feed*  Prior feed*  Stomach wash*  Days of CPAP  Days of Mechanical ventilation  Days of IV cannula  Days of PICC line  Days of Umbilical catheter  Days of Parenteral nutrition | Feeding difficulty  Vomiting  Diarrhoea*  Abdominal distension  Breathing difficulty  Apnoea  Respiratory rate > 60/min  Severe chest indrawing  Cyanosis  Grunting  Fever  Cold to touch  Temperature > 37.5 C  Temperature < 36.5 C  Heart rate > 180/min  Heart rate < 100/min  Capillary refill > 3 sec  Convulsions  Moves only when stimulated  Lethargy  Bulging fontanelle*  Ear discharge  Pus from umbilicus*  Skin pustules  Sepsis screen |

**Nine variables excluded from Boruta feature selection*

**Supplementary figure S2. Correlation matrix of variables**

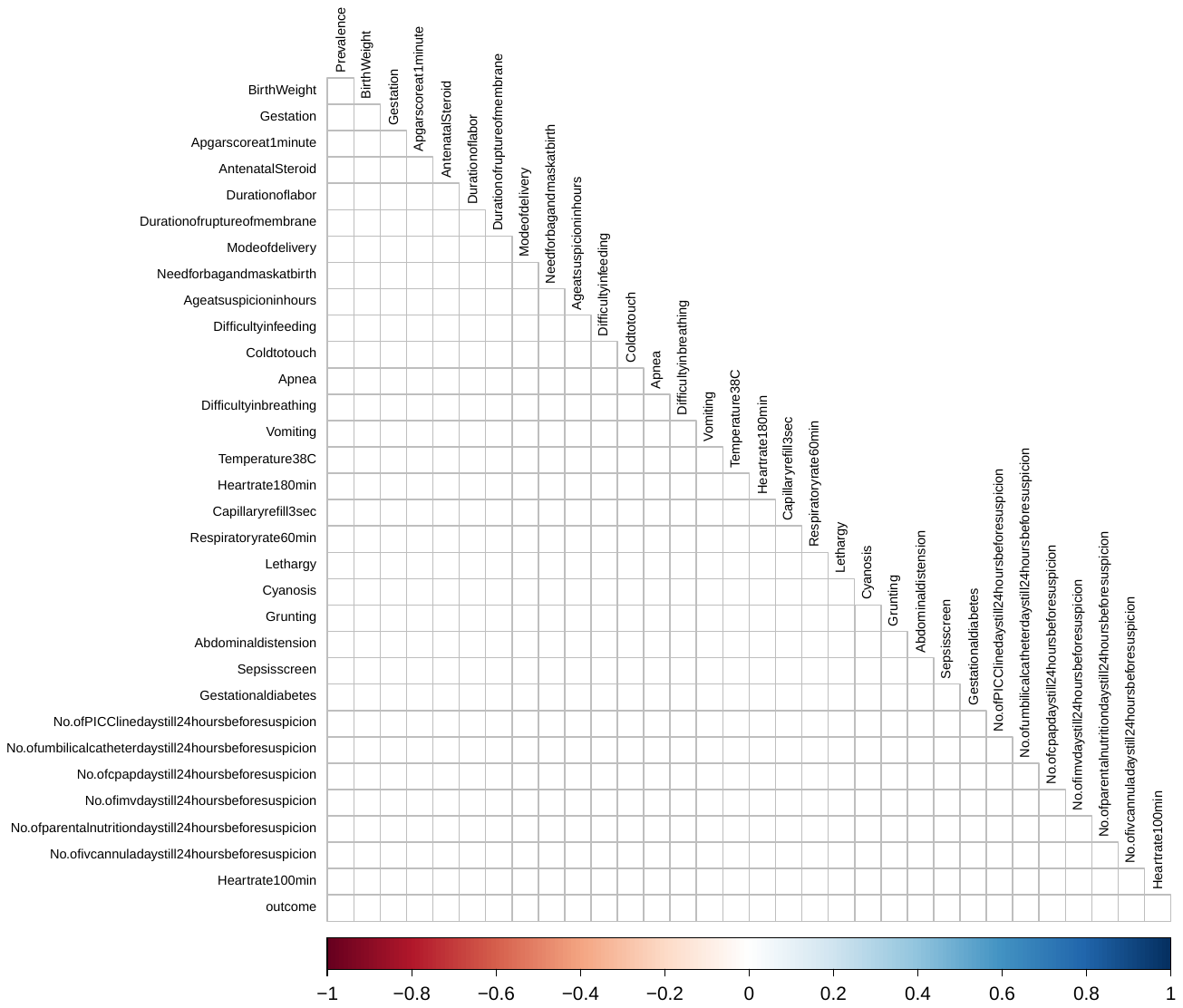

**Supplementary table S4. List of features selected using lasso feature selection**

| **Maternal characteristics** | **Neonatal characteristics** | **Clinical features** |
| --- | --- | --- |
| - Mode of delivery - Gestational diabetes | - Age at suspicion of sepsis (hours)*   ***Care-related variables:****   - Days of mechanical ventilation - Days of intravenous cannula - Days of umbilical catheter | - Diarrhoea - Abdominal distension - Respiratory rate > 60/min - Heart rate > 180/min or   <100/min   - Lethargy - Sepsis screen |

**Expressed as continuous data*

**Supplementary table S5. Comparison of characteristics of the test data and training data during model development phase**

| **Variable** | **Train data**  **(n=2340)** | **Test data**  **(n=584)** |
| --- | --- | --- |
|  | *Maternal characteristics* |  |
| Gestation (weeks), mean (SD) | 35.3 (3.7) | 35.3 (3.8) |
| Maternal fever, n (%) | 266 (11.3) | 67 (11.4) |
| Maternal antibiotics, n (%) | 677 (28.9) | 166 (28.3) |
| No. of vaginal examination*^a^* | 2 (2,3) | 2 (2, 3) |
| Duration of labor (hours) *^a^* | 7 (2,10) | 7 (4, 12) |
| Duration of rupture of membranes (hours) *^a^* | 9 (0, 26) | 9 (0, 26) |
| Caesarean section, n (%) | 743 (31.7) | 192 (32.8) |
| Meconium-stained liquor, n (%) | 517 (22) | 145 (24.7) |
| Foul smelling liquor, n (%) | 114 (4.8) | 24 (4.1) |
| Maternal age (years) *^a^* | 24 (22, 27) | 24 (22, 27) |
| Multiple births, n (%) | 226 (9.6) | 61 (10.4) |
| No. of antenatal visits*^a^* | 6 (4, 9) | 6 (4, 8) |
| Primigravida, n (%) | 1200 (51.2) | 297 (50.7) |
| Maternal urinary tract infection, n (%) | 206 (8.8) | 44 (7.5) |
| Maternal medical/surgical illness, n (%) | 266 (11.3) | 67 (11.4) |
| Antenatal steroids*, n (%) | 644 (27.5) | 162 (27.6) |
| Obstetric problem, n (%)   - Hypertension - Anemia - Gestational diabetes | 299 (12.7)  254 (10.8)  49 (2.0) | 85 (14.5)  70 (11.9)  12 (2.0) |
| Centre, n   - Site 1 - Site 2 - Site 3 | 135  203  2002 | 26  53  506 |
| *Neonatal characteristics* | | |
| Birth weight (g), mean (SD) | 2112 (754) | 2096 (761) |
| Male gender, n (%) | 1705 (58.3) | 337 (57.6) |
| Apgar at 1 min*^a^* | 7 (5, 8) | 7 (4, 8) |
| Apgar at 5 min*^a^* | 8 (7, 9) |  |
| Need for bag and mask, n (%) | 959 (32.8) | 206 (35.2) |
| Need for chest compressions, n (%) | 62 (2.1) | 10 (1.7) |
| Major malformation, n (%) | 165 (5.6) | 31 (5.2) |

*SD: Standard deviation*

*^a^ Data expressed as median with interquartile range*

**Supplementary figure S3. Performance of the RF model at various probability thresholds from 0 to 0.5**

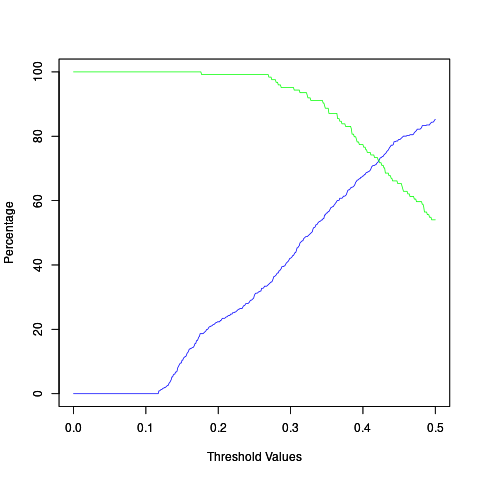

*The green line indicates values for sensitivity (percentage) and blue line indicates values of specificity for various probability thresholds*

**Supplementary table S6. Performance metrics of machine learning model using Boruta feature selection**

| **ML model** | **Sensitivity** | **Specificity** | **PPV** | **NPV** |
| --- | --- | --- | --- | --- |
| RF (0.33) | **90.2** | **40.6** | **25.3** | **95.1** |
| GBM (0.36) | 90.1 | 35.7 | 23.8 | 93.9 |
| XGBOOST (0.25) | 90.3 | 35.3 | 23.9 | 93.8 |
| SVM (0.20) | 82.2 | 35.2 | 22.2 | 89.8 |
| Ensemble (0.32) | 65.3 | 25.1 | 16.4 | 76.4 |

*The numbers in parentheses denote the probability threshold. The numbers in each cell expressed as percentage. The boldfaced numbers indicate the best metrics of each model.*

*RF: Random forest, GBM: Gradient boosting machine, XGBOOST: Extreme gradient boosting, SVM: Support vector machine, AUROC: Area under Receiver Operating Characteristics, PPV: Positive predictive value, NPV: Negative predictive value*

**Supplementary table S7. 2x2 table depicting the performance of the Random Forest model in the development phase using test data**

|  | ***Test data (20%)*** | | |
| --- | --- | --- | --- |
|  | **Diagnosis** | |  |
| **Prediction** | Culture-positive  sepsis | No sepsis | Total |
| Sepsis | 96 | 283 | 380 |
| No Sepsis | 10 | 194 | 204 |
| **Total** | 107 | 477 | 584 |

**Supplementary table S8. Performance of models developed using lasso feature selection**

| **ML model** | **Sensitivity** | **Specificity** | **PPV** | **NPV** |
| --- | --- | --- | --- | --- |
| RF (0.29) | 90.2 | 23.9 | 20.9 | 91.2 |
| GBM (0.36) | 90.3 | 22.4 | 20.5 | 91.4 |
| XGBOOST (0.25) | 90.3 | 25.1 | 21.1 | 92.3 |
| SVM (0.2) | 82.1 | 21.5 | 19.0 | 84.4 |
| Ensemble (0.32) | 65.4 | 22.0 | 15.8 | 73.9 |

*The numbers in parentheses denote the probability threshold. The numbers in each cell expressed as percentage. The boldfaced numbers indicate the best metrics of each model.*

*RF: Random forest, GBM: Gradient boosting machine, XGBOOST: Extreme gradient boosting, SVM: Support vector machine, AUROC: Area under Receiver Operating Characteristics, PPV: Positive predictive value, NPV: Negative predictive value*

**Supplementary table S9. Baseline characteristics of neonates enrolled in the validation phase**

| **Characteristic** | **n=147** |
| --- | --- |
| Birth weight (g), mean (SD) | 1570 (703) |
| Gestation (completed weeks), mean (SD)   - - 37 weeks or more, n (%) - - 34-36 weeks, n (%) - - 28-33 weeks, n (%)   - < 28 weeks, n (%) | 31 (4)  20 (13.6)  25 (17)  77 (52.4)  25 (17) |
| Males, n (%) | 90 (61.2) |
| Apgar at 1 min***^a^*** | 7 (2-8) |
| No. of antenatal visits ***^a^*** | 4 (4-5) |
| Primigravida, n (%) | 51 (34.7) |
| No. of vaginal examination during labor ***^a^*** | 1 (0-2) |
| Duration of rupture of membranes (hours)***^a^*** | 3 (0-36) |
| Vaginal delivery, n (%) | 67 (45.6) |
| Maternal fever, n (%) | 11 (7.5) |
| Meconium-stained liquor, n (%) | 10 (6.8) |
| Risk factor based antibiotic therapy, n (%) | 35 (23.9) |
| Age at suspicion (hours)***^a^*** | 10 (0-28) |
| Antibiotics received at suspicion, n (%) | 110 (75.3) |
| Duration of antibiotic therapy (days)***^a^*** | 5 (4-7) |
| *Clinical features*  Abdominal distension, n (%)  Vomiting, n (%)  Difficulty in breathing, n (%)  Tachypnoea, n (%)  Cyanosis, n (%)  Apnoea, n (%)  Fever, n (%)  Hypothermia, n (%)  Tachycardia, n (%)  Bradycardia, n (%)  Delayed capillary refill, n (%)  Lethargy, n (%)  Convulsions, n (%)  Diarrhoea, n (%)  Skin pustules, n (%) | 15 (10.2)  12 (8.2)  62 (42.2)  61 (41.5)  76 (51.7)  33 (22.5)  5 (3.4)  10 (6.8)  18 (12.2)  5 (3.4)  44 (29.9)  7 (4.8)  2 (1.4)  0  5 (3.4) |
| *Care related factors*  CPAP, n (%)  IMV, n (%)  IV cannula, n (%)  PICC line, n (%)  Umbilical catheter, n (%)  Arterial line, n (%)  Parenteral nutrition, n (%) | 19 (12.9)  16 (10.9)  26 (17.7)  3 (2.0)  23 (15.6)  0  14 (9.5) |
| *Sepsis screen*  Positive, n (%)  Negative, n (%)  Not done, n (%) | 22 (15.0)  89 (60.5)  36 (24.5) |

*SD: Standard deviation* ***^a^*** *Data expressed as median with interquartile range*

**Supplementary table S10. 2x2 table depicting the performance of the Random Forest model in the validation phase**

|  | **Diagnosis** | |  |
| --- | --- | --- | --- |
| **Prediction** | Culture-positive  sepsis | No sepsis | Total |
| Sepsis | 24 | 76 | 100 |
| No Sepsis | 2 | 45 | 47 |
| **Total** | 26 | 121 | 147 |

**Supplementary table S11. 2x2 table depicting the performance of the Random Forest model in the validation phase**

|  | **Diagnosis** | |  |
| --- | --- | --- | --- |
| **Prediction** | Culture-positive  sepsis | No sepsis | Total |
| Sepsis | 3 | 21 | 24 |
| No Sepsis | 0 | 23 | 23 |
| **Total** | 3 | 44 | 47 |

**Supplementary panel S2. Case-based scenarios from the validation phase**

***Case A***

- Late preterm, low birth weight
- *Perinatal risk factors:*
- Prolonged rupture of membranes
- Four per vaginal examinations during labor

The neonate was asymptomatic.

***Remarks:***

The model correctly classified the episode as No Sepsis. (see figure below) Antibiotics would have been safely avoided in this neonate using our model. The red bars indicate the factors that contradict the predicted output and blue bars indicate those that support the output.

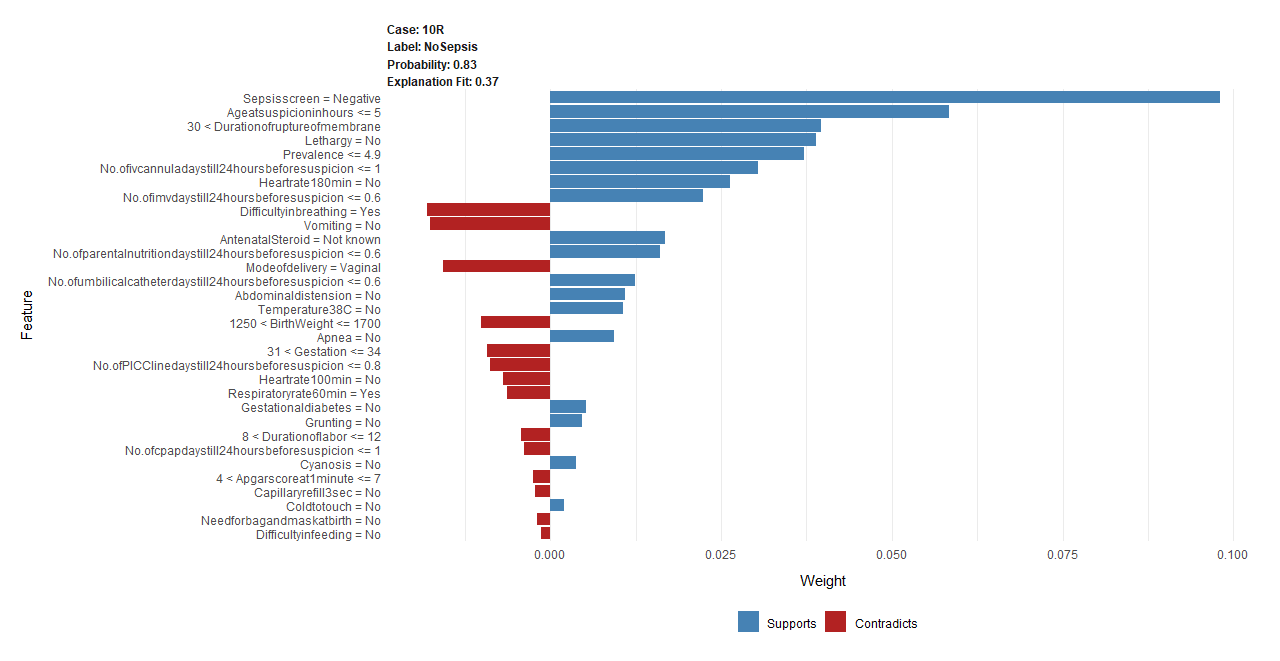

***Case B***

- Extreme preterm, extremely low birth weight
- At 36 hours: Respiratory distress, hypothermia, delayed capillary refill
- Sepsis screen negative

***Remarks:***

The outcome was wrongly predicted as Sepsis (see figure below) due to weight <1250g, gestation <31 weeks, clinical signs, and presence of umbilical catheter for at-least 24 hours. The actual diagnosis was No Sepsis.

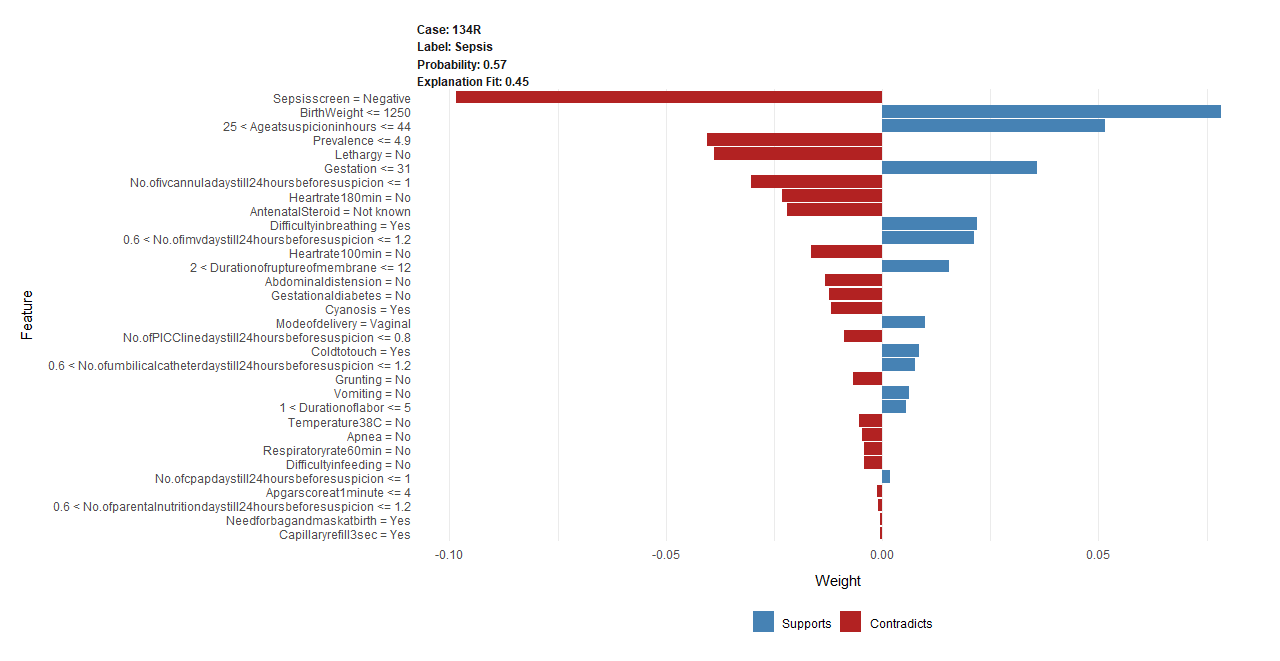

***Case C***

Extreme preterm, extremely low birth weight, no perinatal risk factors

- At 44 hours: Worsening hypoxia, recurrent apneas
- Sepsis screen negative

***Remarks:***

The culture was positive, and the model would have appropriately labelled this episode as Sepsis (see figure below) and led to antibiotic initiation without delay. The sepsis screen was notably negative

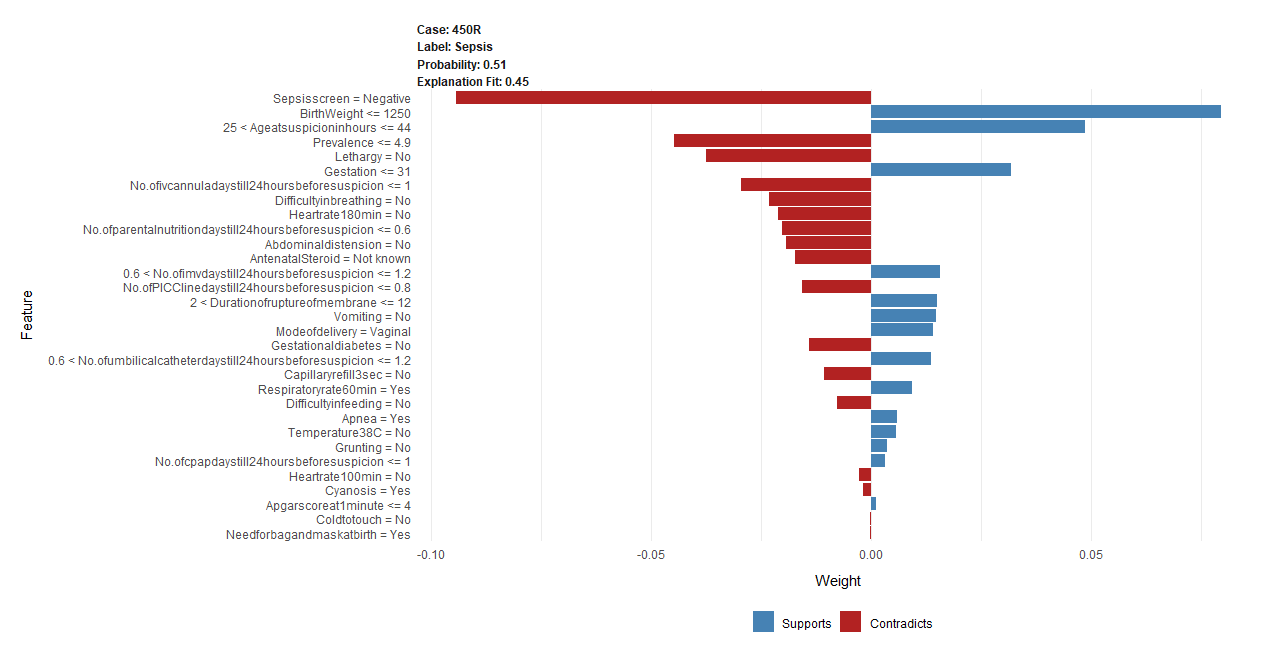

**Supplementary panel S2. Case-based scenarios from the validation phase (continued)**
